# Interictal intracranial EEG asymmetry lateralizes temporal lobe epilepsy

**DOI:** 10.1101/2023.12.13.23299907

**Authors:** Erin C. Conrad, Alfredo Lucas, William K.S. Ojemann, Carlos A. Aguila, Marissa Mojena, Joshua J. LaRocque, Akash R. Pattnaik, Ryan Gallagher, Adam Greenblatt, Ashley Tranquille, Alexandra Parashos, Ezequiel Gleichgerrcht, Leonardo Bonilha, Brian Litt, Saurabh Sinha, Lyle Ungar, Kathryn A. Davis

**Affiliations:** Department of Neurology, Perelman School of Medicine, University of Pennsylvania, Philadelphia, PA 19104, USA; Center for Neuroengineering and Therapeutics, University of Pennsylvania, Philadelphia, PA 19104, USA; Department of Bioengineering, School of Engineering & Applied Sciences, University of Pennsylvania, Philadelphia, PA 19104, USA; Perelman School of Medicine, University of Pennsylvania, Philadelphia, PA 19104, USA; Department of Neurology, Washington University in St. Louis, St. Louis, MO 63110, USA; Department of Neurology, Medical University of South Carolina, Charleston, SC 29425, USA; Department of Neurology, Emory University, Atlanta, GA 30325, USA; Department of Computer and Information Science, University of Pennsylvania, Philadelphia, PA 19104, USA

**Keywords:** EEG, intracranial EEG, temporal lobe epilepsy, interictal EEG

## Abstract

Patients with drug-resistant temporal lobe epilepsy often undergo intracranial EEG recording to capture multiple seizures in order to lateralize the seizure onset zone. This process is associated with morbidity and often ends in postoperative seizure recurrence. Abundant interictal (between-seizure) data is captured during this process, but these data currently play a small role in surgical planning. Our objective was to predict the laterality of the seizure onset zone using interictal (between-seizure) intracranial EEG data in patients with temporal lobe epilepsy. We performed a retrospective cohort study (single-center study for model development; two-center study for model validation). We studied patients with temporal lobe epilepsy undergoing intracranial EEG at the University of Pennsylvania (internal cohort) and the Medical University of South Carolina (external cohort) between 2015 and 2022. We developed a logistic regression model to predict seizure onset zone laterality using interictal EEG. We compared the concordance between the model-predicted seizure onset zone laterality and the side of surgery between patients with good and poor surgical outcomes. 47 patients (30 women; ages 20-69; 20 left-sided, 10 right-sided, and 17 bilateral seizure onsets) were analyzed for model development and internal validation. 19 patients (10 women; ages 23-73; 5 left-sided, 10 right-sided, 4 bilateral) were analyzed for external validation. The internal cohort cross-validated area under the curve for a model trained using spike rates was 0.83 for a model predicting left-sided seizure onset and 0.68 for a model predicting right-sided seizure onset. Balanced accuracies in the external cohort were 79.3% and 78.9% for the left- and right-sided predictions, respectively. The predicted concordance between the laterality of the seizure onset zone and the side of surgery was higher in patients with good surgical outcome. In conclusion, interictal EEG signatures are distinct across seizure onset zone lateralities. Left-sided seizure onsets are easier to distinguish than right-sided onsets. A model trained on spike rates accurately identifies patients with left-sided seizure onset zones and predicts surgical outcome.

## INTRODUCTION

Temporal lobe epilepsy (TLE) is the most common localization of drug-resistant epilepsy in adults ^1^. Determining the laterality of TLE — left, right, or bilateral — is a common clinical question that dictates the type and location of surgery. One standard clinical approach to confirming the localization and laterality of suspected TLE is to implant intracranial electrodes and wait 1-2 weeks for the patient to have multiple seizures ^2^. This process is time-consuming and exposes the patient to morbidity associated with electrode implantation, recurrent seizures, and prolonged hospitalization. Ironically, it may also be too short to determine epilepsy laterality: Data from chronic intracranial EEG recordings reveal that, in patients with bilateral seizure onsets, it often takes several weeks to months to capture the first contralateral seizure ^3^. Even in the setting of weaning medications in the Epilepsy Monitoring Unit, patients with a moderate pretest probability of multifocal seizures may need seven or more seizures to achieve a high posttest probability of unifocal epilepsy, which is more seizures than can often be obtained in a 1-2 week intracranial EEG evaluation ^4^. There is a pressing need to identify biomarkers of epilepsy laterality *that are persistently abnormal*, and thus identifiable during the limited duration of a typical pre-surgical evaluation. Such biomarkers could be used to reduce morbidity, shorten the hospital length of stay, and improve surgical planning.

During the course of a typical intracranial EEG evaluation, massive amounts of *interictal data* — in between the seizures — are obtained. However, these data are clinically underutilized because, up until now, we have lacked methods to interpret such large quantities of data. In this study, we hypothesized that features of the interictal EEG differ between patients with left-sided, right-sided, and bilateral TLE. We studied interictal intracranial EEG data from patients with drug-resistant TLE. We calculated several interictal EEG features and compared these features between patients with left, right, and bilateral TLE. Next, in order to understand if lateralizing interictal features exist across modalities, we studied functional MRI (fMRI) connectivity in a separate group of patients with TLE. Finally, we developed a machine learning algorithm to predict the clinician-defined seizure onset zone laterality using interictal data, and then validated this model across patients from two epilepsy centers.

## MATERIALS AND METHODS

### Patient selection

This retrospective study was approved by the Institutional Review Boards of the Hospital of the University of Pennsylvania (HUP) and Medical University of South Carolina (MUSC). We analyzed patients with drug-resistant epilepsy who underwent intracranial EEG recording as part of surgical evaluation from 2015-2022. Inclusion criteria were 1) clinical determination of TLE, and 2) bilateral electrode coverage of the temporal lobes. Details on electrode configurations, recording, and determination of seizure onset zone and surgical outcome are in the Supplemental Methods. Seizure times and the anatomical seizure onset localization were identified by a board-certified epileptologist reviewing the EEG record for clinical purposes, and confirmed in a clinical case conference consisting of multiple epileptologists. For patients who underwent resection or ablation, we measured surgical outcome at one year post-surgery using both the ILAE and Engel classification scales ^5,6^.

### Intracranial EEG pre-processing, feature selection, and asymmetry index calculation

A full description of temporal and spatial sampling is in the Supplemental Methods. Briefly, we selected a continuous 12-hour period of EEG data and applied the SleepSEEG algorithm ^7^ to determine sleep stage. We divided the 12-hour EEG period into 72 10-minute segments, from which we randomly chose one-minute segments. We removed non-temporal lobe electrode contacts and those contacts lacking a contralateral pair (Fig. 1A) because analyzing symmetric electrode pairs limits the confounder of inter-contact distance affecting functional connectivity measurements ^8,9^. We identified and eliminated channels with artifacts ^10^ and employed three different referencing methods: machine reference, common average reference, and bipolar reference (referencing was performed after removing non-symmetric and non-temporal lobe contacts). We applied a 60 Hz notch filter and a 0.5-80 Hz bandpass filter.

**Figure 1.**
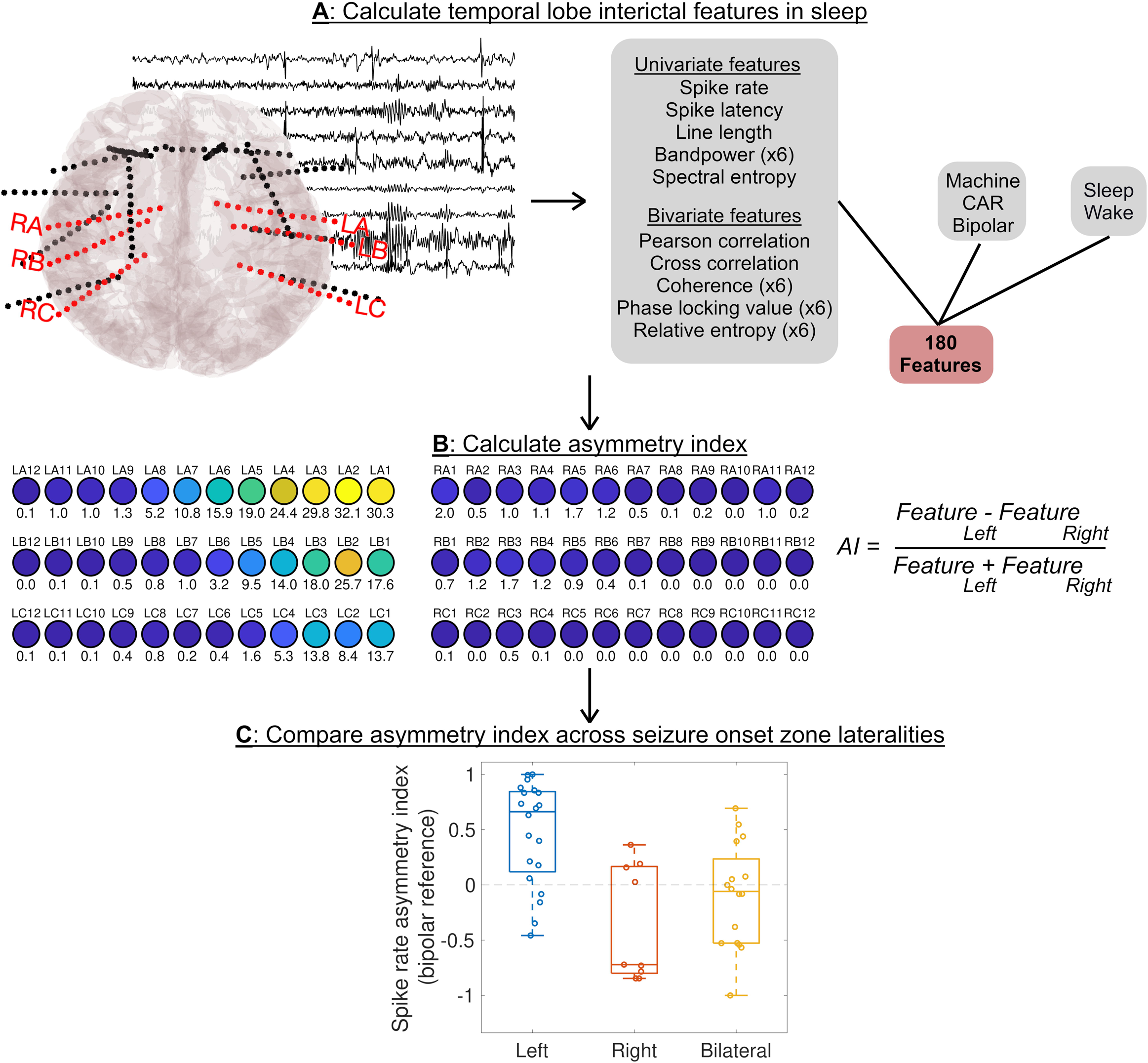
Methods to measure interictal IEEG asymmetry index (AI). **A:** We identified electrodes targeting the bilateral temporal structures, and calculated several univariate and bivariate (connectivity) features for three choices of reference (machine, common average, and bipolar) and in both sleep and wake, resulting in a total of 180 features. **B:** We calculated the asymmetry index for each feature. **C:** We compared the asymmetry index across patients with different seizure onset zone lateralities.

We calculated several features, chosen based on their prior use in the literature for identifying the seizure onset zone (Fig. 1A). The Supplemental Materials fully describe the calculation of each feature. Briefly, these features included spike rates, univariate features such as bandpower and line length, and several bivariate features representing functional connectivity. Some features were calculated only over the broadband signal, and some were also calculated over the EEG signal filtered to five canonical frequencies. We separately examined features in sleep and wake given prior work suggesting that sleep-wake rhythms of seizure-risk differ between left and right TLE ^10^, and given work suggesting that interictal features in sleep better localize seizure generators than features in wakefulness ^6,9^.

We next calculated the *asymmetry index* (AI) for each feature, defined as:

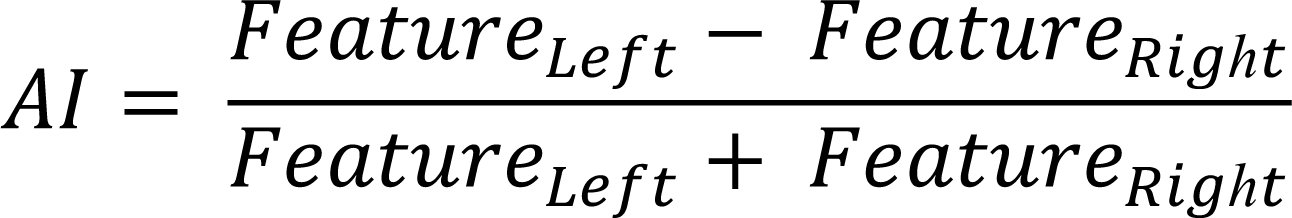

Where *Feature_Left_* is the mean feature across left temporal lobe contacts, and *Feature_Right_* is the mean feature across right temporal lobe contacts. A positive AI implies higher values of the feature on the left, and a negative AI implies higher values of the feature on the right.

### fMRI analysis

To determine if interictal asymmetries from non-EEG modalities also lateralize TLE, we studied fMRI connectivity using the dataset and processing techniques from a recent paper published by our group ^11^. Subjects were 62 patients at HUP with drug-resistant TLE, six of whom overlapped with those in our intracranial EEG cohort. We calculated the average BOLD time series for each voxel within each parcel of the Brainnetome atlas ^12^ and created a functional connectivity matrix by computing the absolute value of the Pearson correlation between the time series in each pair of parcels. We identified Brainnetome parcels belonging to temporal lobe gray matter structures (Fig. 2C, Table S2), and measured the average left and right temporal connectivity. We defined *AI* as:

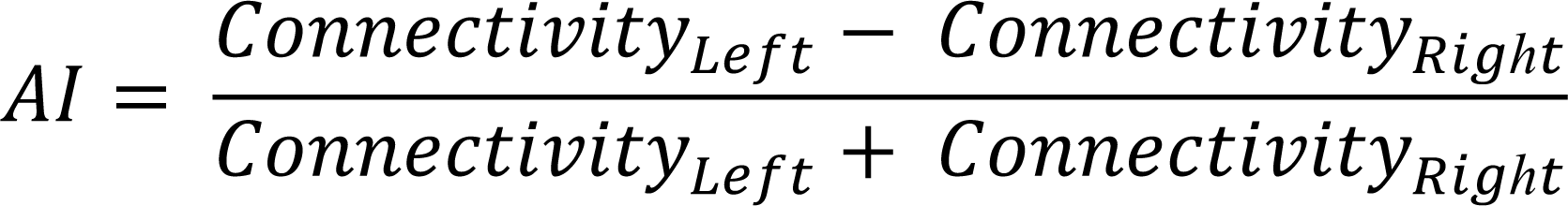

**Figure 2.**
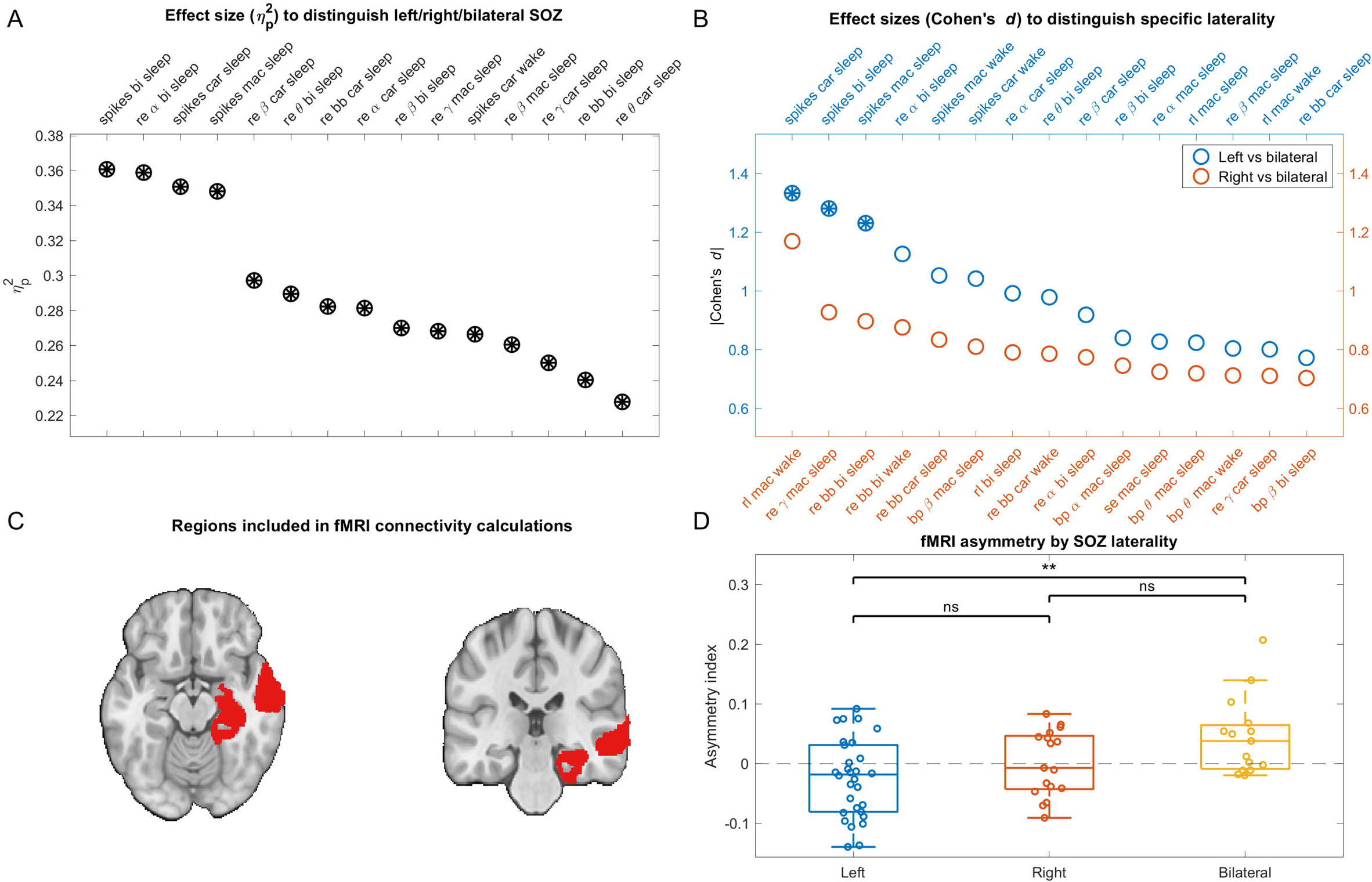
Comparison of IEEG and fMRI asymmetry index (AI) across seizure onset zone (SOZ) lateralities. **A:** Effect sizes (η^2^) at distinguishing patients with different SOZ lateralities for the 15 features with the highest effect sizes. Circles filled with asterisks (all in this plot) represent features that significantly distinguished SOZ lateralities (ANOVA with Benjamini-Hochberg false discovery rate correction). **B:** Effect sizes (absolute value of Cohen’s *d*) at distinguishing SOZ lateralities for the 15 features with the highest effect sizes at distinguishing left from bilateral SOZs and right from bilateral SOZs, respectively. Note that in this analysis, as opposed to that for Figure 3, we separated left from bilateral and right from bilateral, rather than left from right/bilateral and right from left/bilateral. We did this to achieve distinct groups to better understand the separate univariate features that distinguished left and right TLE. **C:** Regions (Brainnetome parcels) included in fMRI connectivity calculations (shown for the left temporal lobe only). **D:** fMRI AI by SOZ laterality. Both IEEG and fMRI AI distinguished left from bilateral SOZs more easily than right from bilateral SOZs. bi = bipolar reference; car = common average reference; mac = machine reference. spikes = spike rates; re = relative entropy; rl = spike recruitment latency (a measure of spike timing); bp = bandpower; se = spectral entropy. Greek letters indicate canonical frequency bands. iEEG Analyses were performed in the HUP cohort to preserve the MUSC cohort data for external model validation.

Where *Connectivity_Left_* is the mean connectivity in the left temporal lobe, and *Connectivity_Right_* is the mean connectivity in the right temporal lobe.

### Machine learning to predict SOZ laterality

Detailed steps are in the Supplemental Methods. Briefly, to separately examine the ease of predicting left versus right SOZ laterality, we built two classifiers: one predicting left-sided SOZ (as opposed to either right *or* bilateral), and one predicting right-sided SOZ (as opposed to left or bilateral), both using interictal EEG AI values as features. Missing features were estimated by median imputation. For each classifier, we performed principal component analysis (PCA) on the training data, retaining enough components to explain 95% of the variance in the features. We performed LASSO logistic regression (with λ equal to 1/N, where N is the training sample size) on the preserved principal components. For each classifier, we performed internal validation on the HUP dataset using leave-one-patient-out cross-validation, and then external validation on the MUSC dataset, trained on the full HUP dataset. To estimate feature importance, we multiplied the PCA transformation matrix by the classifier feature weights in order to derive standardized weights for the original set of AI features ^13^.

We next developed two single-feature models: The first included only mean spike rate AI. This model was developed to test whether spike rate asymmetry alone could successfully lateralize TLE. The second single-feature model included *binarized* mean spike rate AI, with the input being 1 if the spike rate AI was positive and 0 if negative. This model approximated a qualitative clinical approach of considering which side has more spikes, rather than the quantitative difference between sides. We primarily analyzed spikes detected in common average reference, with secondary analyses of bipolar and machine references to test the sensitivity of the results to this choice. In order to have a single feature, we also chose to only study spikes in sleep given work suggesting that interictal features in sleep better localize seizure generators than features in wakefulness ^14,15^.

Clinician determinations of SOZ laterality may be inaccurate. Given our hypothesis that interictal features reveal seizure generators, we predicted that our models would perform better in those with good surgical outcomes. To test this, we examined the subset of patients who underwent surgical resection or ablation targeting the temporal lobe and had at least one year of surgical follow up. For each patient, we identified the side of surgery, and we measured the spike rate model-predicted probability of SOZ laterality on that side. We selected the model (left vs. right/bilateral, or right vs. left/bilateral) corresponding to the side of surgery. We compared model probabilities between patients with a good surgical outcome and those with a poor surgical outcome.

### Statistical analysis

For univariate analyses, we report mean and standard deviation (SD). To compare two paired or independent groups, we report *t*-tests (alpha = 0.05) and Cohen *d* for effect size. To compare more than two groups, we report ANOVA tests and η^2^ for effect size. Analyses were performed in Matlab R2022a (Mathworks).

### Data and code availability

Raw EEG data is available on ieeg.org. All code used to perform analyses, along with an intermediate dataset containing electrode contact-level features, is publicly available on https://github.com/penn-cnt/cnt_tle_laterality/. The online calculator to predict SOZ laterality given temporal lobe spike rates is available on https://penn-cnt.github.io/epilepsy_lateralization/.

## RESULTS

We examined 47 patients at HUP for model development and internal validation, and 19 patients at MUSC for external model validation (Table 1). We visually validated a random sample of 50 automated spike detections from each patient (bipolar montage). The median (IQR) percentage of automatically-detected spikes determined to be true spikes was 90.0% (82.0%-95.5%) for HUP and 90.0% (76.0%-94.0%) for MUSC.

**Table 1.** Clinical information. The first column shows the clinical variables, the second column shows the data for the cohort undergoing intracranial EEG (IEEG) at HUP, the third column shows data for the cohort undergoing IEEG at MUSC used for external model validation, and the fourth column shows data for the cohort undergoing fMRI at HUP. The “Bilateral or discordant pre-implant hypotheses” variable indicates the number of patients for whom one of the leading two pre-implant localization hypotheses was bilateral, or for whom the leading two pre-implant hypotheses had discordant lateralities (e.g., the top hypothesis was left temporal lobe epilepsy and the second hypothesis was right temporal lobe epilepsy). Patients included in the outcome analysis are those who underwent resection or ablation and had at least one year of follow up.

|  | HUP IEEG | MUSC IEEG | HUP fMRI |
| --- | --- | --- | --- |
| Total: N | 47 | 19 | 62 |
| Female: N (%) | 30 (63.8%) | 10 (52.6%) | 33 (53.2%) |
| Age at onset in years: median (range) | 16.0 (0.9-59.0) | 25.0 (8.0-59.0) | 20.0 (0.3-61.0) |
| Age at implant in years: median (range) | 38.0 (20.0-69.0) | 36.5 (23.0-73.0) | 35.3 (18.9-70.4) |
| Bilateral or discordant pre-implant hypotheses: N (%) | 33 (70.2%) | 17 (89.5%) | NA |
| Symmetric temporal-targeted contacts: median (range) | 44.0 (8.0-72.0) | 60.0 (20.0-60.0) | NA |
| SOZ lateralization (clinician determination) |  |  |  |
| Left: N (%) | 20 (42.6%) | 5 (26.3%) | 30 (48.4%) |
| Right: N (%) | 10 (21.3%) | 10 (52.6%) | 17 (27.4%) |
| Bilateral: N (%) | 17 (36.2%) | 4 (21.1%) | 15 (24.2%) |
| Surgery performed |  |  |  |
| Resection: N (%) | 8 (17.0%) | 5 (26.3%) | 10 (16.1%) |
| Ablation: N (%) | 17 (36.2%) | 2 (10.5%) | 16 (25.8%) |
| Device: N (%) | 14 (29.8%) | 6 (31.6%) | 9 (14.5%) |
| Engel 1 year outcome | N = 24 with outcomes | N = 2 with outcomes | N = 13 with outcomes |
| Median (range) | 1.0 (1.0-4.0) | 1.0 (1.0-1.0) | 1.0 (1.0-3.0) |
| ILAE 1 year outcome | N = 24 with outcomes | N = 2 with outcomes | N = 13 with outcomes |
| Median (range) | 2.0 (1.0-5.0) | 1.0 (1.0-1.0) | 3.0 (1.0-5.0) |

### Asymmetries in spike rates and relative entropy are distinct across SOZ lateralities

We compared interictal EEG feature AIs between patients with left-sided, right-sided, and bilateral SOZs. We ranked features in descending order by effect size (η^2^) at separating the three SOZ lateralities (Fig. 2A). The top-ranked AI features involved spike rates and relative entropy, and were predominantly in sleep. We next compared the set of features that best distinguished left from bilateral SOZs versus right from bilateral SOZs (Fig. 2B). Spike features significantly distinguished left from bilateral SOZs. For distinguishing right-sided SOZ, several other features performed best (though none were significant after correcting for the false discovery rate). This suggests that right-sided SOZs are harder to distinguish than left-sided SOZs in our dataset. Several interictal features were highly correlated, including spikes and relative entropy (see Fig. S1 and Supplemental Results). We performed a secondary analysis in which we excluded 15 unilateral patients who did not undergo surgery or who had one-year Engel outcomes >1. We observed similar trends in this smaller patient cohort (Supplemental Results, Fig. S2).

### fMRI BOLD connectivity AI also distinguishes SOZ lateralities

We next asked if differences in interictal connectivity between TLE lateralities existed across modalities beyond just EEG. We tested whether fMRI connectivity asymmetry similarly distinguished left, right, and bilateral SOZs. Fig. 2C shows the left temporal Brainnetome atlas regions included for fMRI connectivity analysis. There was a significant difference in fMRI connectivity AI between TLE lateralities (ANOVA: F(2,59) = 6.1, *p* = 0.004, η^2^ = 0.17, Fig. 2D). Only the difference between the left and bilateral SOZ group was significant after correcting for multiple comparisons (*p* = 0.002). Similar to the result for interictal EEG data, this suggests that fMRI temporal lobe connectivity AI distinguishes patients with left from bilateral SOZs, but cannot distinguish between patients with right and bilateral SOZs.

### A classifier incorporating interictal EEG features predicts SOZ laterality

We tested whether interictal features predict SOZ laterality in unseen patients. The AUCs of the ROC of the left- and right-sided internal cross-validation models trained on all features were 0.77 and 0.56, respectively (Fig. 3A). A model trained on only spike rates (restricted to CAR reference and sleep in order to have a single feature) achieved higher AUCs (0.83 and 0.68 for the left and right models, respectively (Fig. 3B)). Finally, a model trained only on binary spike rates indicating whether there were more spikes on the left or the right performed poorly (AUC of 0.57 and 0.39, respectively (Fig. 3C)). Spike rates, spike timing, and bandpower were the most important features for both the left- and the right-sided models (Fig. 3D).

**Figure 3.**
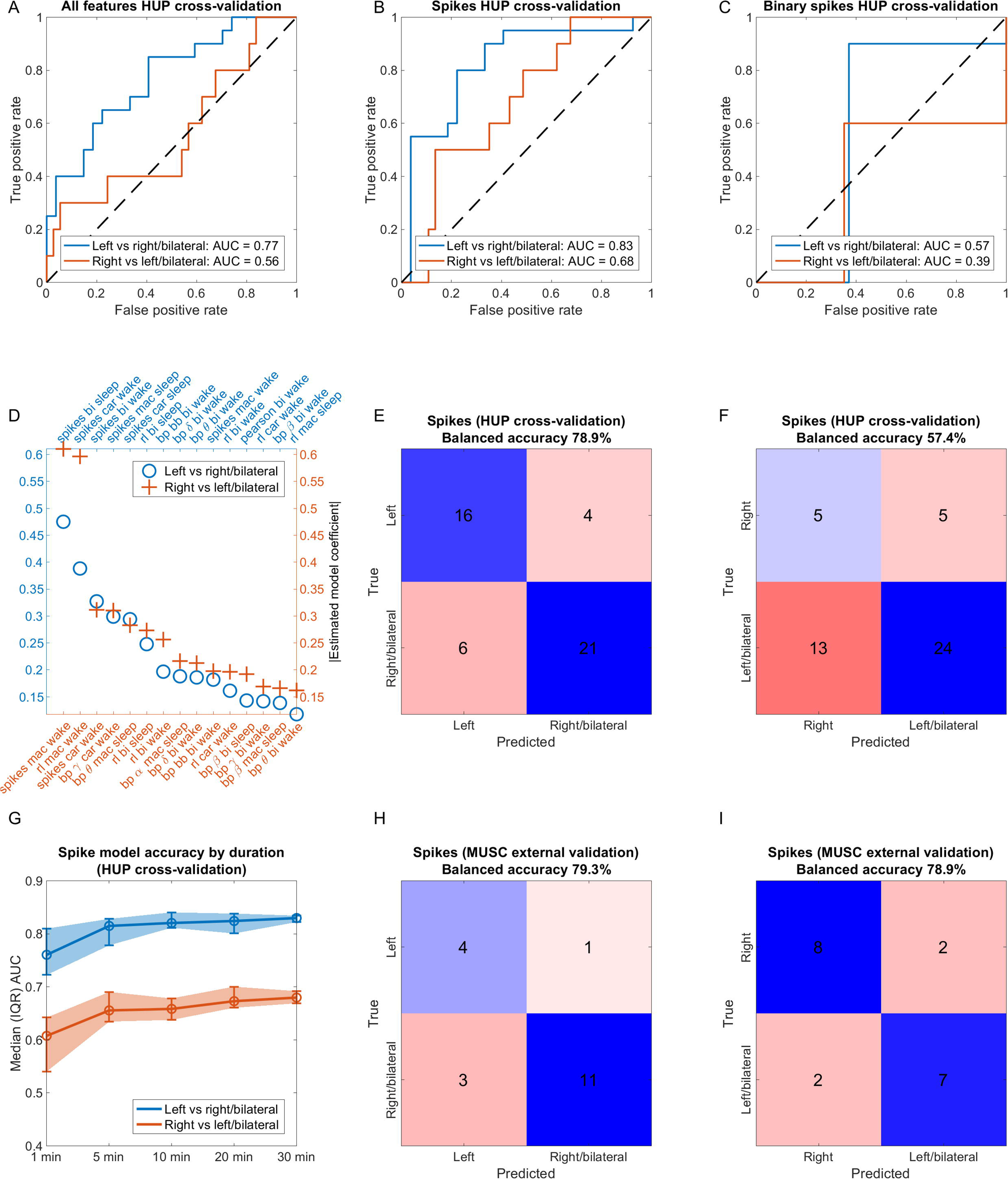
Classifier to distinguish SOZ lateralities using interictal IEEG asymmetry. **A:** Internal cross-validation performance for models trained on all interictal IEEG features. **B:** Internal cross-validation performance for models trained on only spike rate asymmetry index (common average reference). **C:** Internal cross-validation performance for models trained on only spike rate asymmetry index (common average reference), binarized such that 1 = more spikes on the left and 0 = more spikes on the right. **D:** Absolute value of the estimated model coefficients for the full feature set models. mac = machine reference; car = common average reference; bi = bipolar reference. spikes = spike rate; re = relative entropy; bp = bandpower; rl = spike recruitment latency (a measure of spike timing). Greek letters denote canonical frequency bands. **E and F:** Confusion matrices (internal cross-validation) for spike-rate only models (from **B**) at the optimal model operating point for the models predicting left vs. right/bilateral SOZ and right vs. left/bilateral SOZ, respectively. **G:** Model areas under the curve (AUC) for models trained on subsampled durations of spike rate data. **H and I**: Confusion matrices for the spike-rate only models (from **B**) for the models predicting left vs. right/bilateral SOZ and right vs. left/bilateral SOZ, respectively, applied to the external testing set.

We further probed the accuracy of the spike-rate only model. Confusion matrices for the left- and right-sided models at the optimal operating points are shown in Figs. 3E and 3F. The balanced accuracy was 78.9% for the model predicting left vs. right/bilateral SOZ, and 57.4% for the model predicting right vs. left/bilateral SOZ. Model accuracies rise quickly with duration sampled, achieving an accuracy similar to the full-duration models with 5 minutes of sampling (Fig. 3G). Finally, we tested how the spike-only models performed in the external MUSC dataset. The balanced accuracies were 79.3% and 78.9% for the left-sided and right-sided models, respectively (Fig. 3H and 3I).

These results suggest that models using only spike rate asymmetry accurately distinguished left from right or bilateral SOZs in both internal cross-validation and in a separate institution’s test dataset. However, although right-sided SOZs could be distinguished from left/bilateral SOZs in the external dataset set, they were not well-classified in the internal validation dataset. Results were similar, although with higher AUCs across all models, when we restricted analysis of unilateral HUP patients to be those with Engel 1 surgical outcomes to build and internally validate the SOZ laterality classifier (Fig. S3). Results were also similar when we used spikes detected in bipolar and machine references to build the SOZ laterality classifier (Fig. S4 and S5).

### Concordance between spike-predicted laterality and surgical laterality is higher for patients with good surgical outcomes

18 of 26 (69.2%) patients had good one-year Engel outcomes (Engel I), and 8 of 26 (30.8%) had poor Engel outcomes (Engel 2+) (Fig. 4A). 17 of 26 (65.4%) patients had good one-year ILAE outcomes (ILAE 1-2), and 9 of 26 (34.6%) had poor ILAE outcomes (ILAE 3+) (Fig. 4D). The means of the numerical portions of the outcome scales were similar for patients who underwent left versus right-sided surgeries (Fig. 4B and 4E; Engel: *t*(24) = -0.0, *p* = 0.98; ILAE: *t*(24) = 1.0, *p* = 0.31). Left-sided surgeries were disproportionately ablations (12 ablations vs 3 resections), and right-sided surgeries were more often resections (4 ablations vs 7 resections). We hypothesized that patients with a good surgical outcome would have a higher modeled probability of SOZ laterality concordant with the side of surgery. We identified the spike rate model corresponding to the side of surgery. Mean concordant model probability was significantly higher in patients with good Engel outcomes (mean (SD) 0.64 (0.16)) than in patients with poor Engel outcomes (0.48 (0.19)) (*t*(24) = 2.2, *p* = 0.037) (Fig. 4C), and in patients with good ILAE outcomes (0.65 (0.15)) than in patients with poor ILAE outcomes (0.47 (0.18)) (*t*(24) = 2.6, *p* = 0.014) (Fig. 4F). Together, these results suggest that a model trained to predict the SOZ using spike rate asymmetry also predicts surgical outcome. Results were similar when we used spikes detected in bipolar and machine references (Fig. S6 and S7).

**Figure 4.**
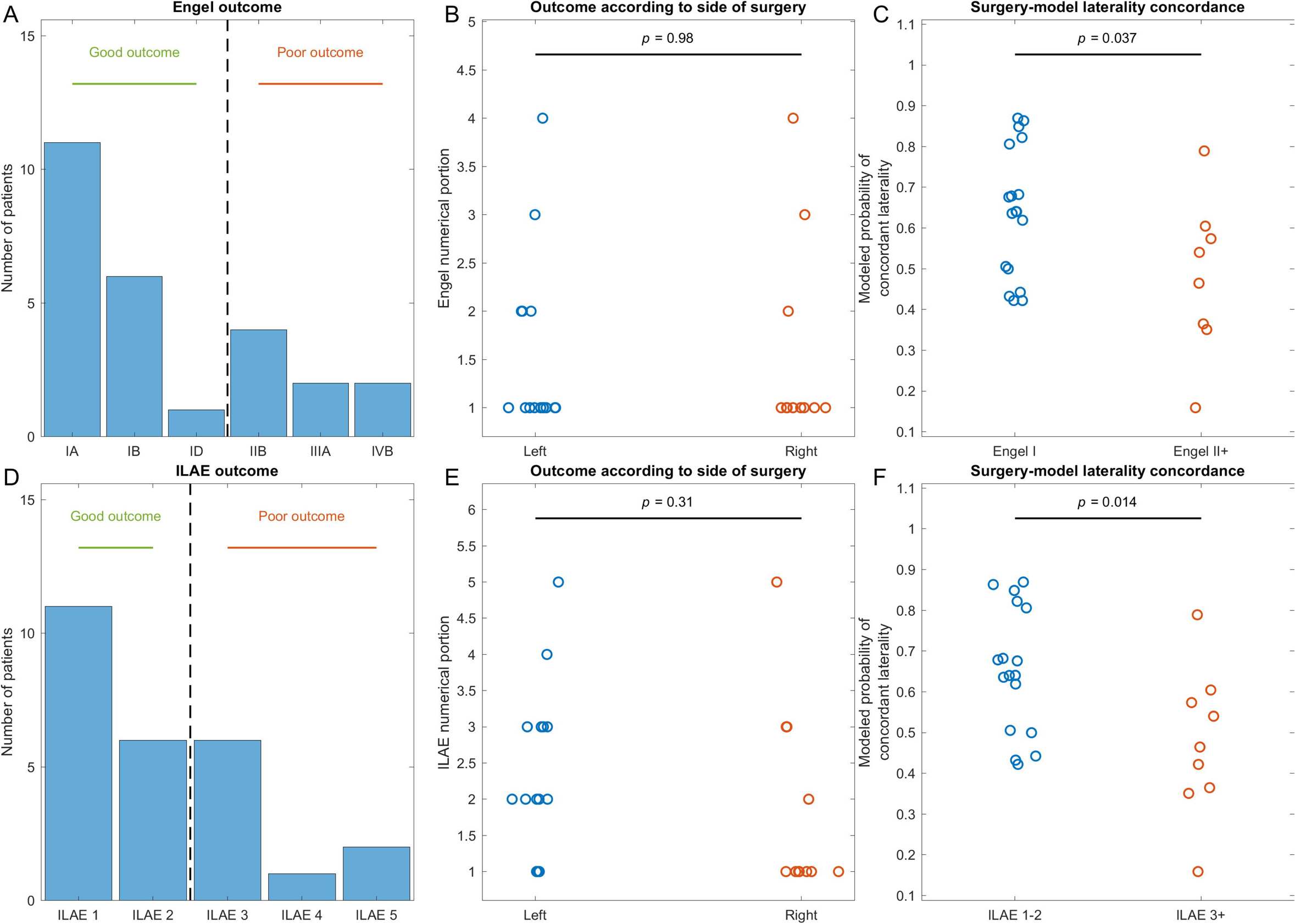
Surgical outcome prediction. **A and D:** Distribution of one-year surgical outcomes using the Engel and ILAE classification schemes, respectively. **B and E**: The numerical portion of the Engel and ILAE surgical outcome score, respectively, according to the side of surgery. **C and F**: The modeled probability of SOZ laterality concordant with the side of surgery for patients who had a good (Engel 1 or ILAE 1-2) or poor (Engel II+ or ILAE 3+) surgical outcome. Patients who had a good surgical outcome had higher laterality-concordant model probabilities than patients who had a poor surgical outcome.

## DISCUSSION

We compared interictal intracranial EEG features across TLE patients with different SOZ lateralities. Left-sided TLE was easier to distinguish than right-sided TLE, a finding we also observed in a separate interictal modality of fMRI connectivity. A model based on just spike rates predicts SOZ laterality in unseen patients.

### Left-sided SOZs are easier to identify than right-sided SOZs

Our univariate analysis of interictal EEG features (Fig. 2B), our machine learning algorithm (Fig. 3A-C), and our fMRI analysis (Fig. 2D) all found that left-sided SOZs were easier to distinguish than right-sided SOZs in our internal HUP cohort. It is possible that this finding is specific to patients who undergo intracranial EEG recording: Patients with well-localized right-sided TLE are more likely to immediately pursue a temporal lobectomy given lower concern about cognitive impact from resecting the non-dominant hemisphere; thus, patients with right-sided TLE who undergo intracranial EEG implantation may have challenging features to their localization. This hypothesis may also explain the discrepancy in right-sided model performance across HUP and MUSC if these two centers have different approaches to select patients with suspected TLE for intracranial EEG monitoring. This hypothesis was not supported by our finding that patients with left-sided and right-sided TLE had similar surgical outcomes, however this is confounded by the fact that patients with left TLE more often underwent laser ablation, which is known to have lower rates of seizure freedom than temporal lobectomy ^16,17^. An alternate hypothesis is that right TLE may be associated with broader network dysfunction. This hypothesis is conceivable due to differences in language networks ^18,19^, and is supported by functional and structural neuroimaging studies demonstrating more widespread abnormalities in right-sided TLE ^20–24^.

### A multivariate set of interictal features is worse than spikes alone

A model using only spike rates distinguished SOZ lateralities with higher accuracy than a model incorporating many interictal features (Fig. 3A and 3B). We suspect that the better performance of the spike-only model arises from the fact that spikes are the most important feature (Fig. 3D), and the benefit of adding more features is outweighed by the downside of overfitting to this more complex feature set. We included intracranial EEG features that have been reported to localize seizure generators in prior studies ^14,25–29^. Relative entropy, a recently-described metric that compares the distribution of amplitudes between EEG signals on electrode pairs ^14,29^, was the non-spike feature that differed the most across lateralities (Fig. 2A), although it was also strongly correlated with spike rates (Fig. S1A and S1C). Overall, we found no clear lateralizing value in these quantitative features beyond spikes alone.

### Spike rate asymmetry identifies left-sided TLE in both internal and external cohorts

The spike-only model accurately identified left-sided TLE in both the internal cross-validation and external cohorts (Fig. 3E and 3H), suggesting good external validity. The performance of the model trained to identify right-sided TLE was low in the internal cross-validation cohort but high in the external cohort (Fig. 3F and 3I). Part of the discrepancy in performance between the two cohorts may result from the class imbalance between the two sites: HUP had more patients with left-sided TLE and MUSC had more patients with right-sided TLE. Alternatively, it may be related to different approaches to surgical planning in the two centers, as discussed above.

Importantly, clinician designations of SOZ laterality may be incorrect: a patient may have only unilateral seizures during the intracranial evaluation and receive a diagnosis of unilateral TLE, but actually have bilateral TLE ^3^. We found that the concordance between the spike model-predicted SOZ laterality and the side of surgery was associated with good one-year surgical outcomes (Fig. 4). This result suggests that a model incorporating spike rates reveals information about the laterality of the true seizure generators.

Prior studies found that spike rates in scalp EEG lateralize TLE, and in particular that when spikes are mostly unilateral, the side of the spikes usually agrees with the side of the clinician-defined epilepsy ^30–34^. Our study adds to these findings by 1) incorporating interictal intracranial EEG data, 2) providing a quantitative framework to interpret non-unilateral spike rates, and 3) evaluating patients undergoing intracranial EEG, who likely have more challenging lateralization than patients with TLE more generally. A future direction is to understand if quantitative analysis of interictal *scalp* EEG data is as effective as invasive data in these patients, which may help avoid intracranial evaluation in some cases.

### Limitations

Both our internal validation cohort and our external validation cohort have relatively few patients, which limits the external validity. We had only six overlapping patients between our fMRI dataset and our IEEG dataset, which precluded us from assessing the complementary information they could provide in a multimodal approach. Finally, because our machine learning algorithm does not predict SOZ *localization*, we cannot identify patients whose seizure generators were correctly lateralized, but incorrectly localized. This is a particular limitation for those patients who underwent laser ablation, where precisely localizing seizure generators is crucial.

### Clinical translation

As spike rates can be counted manually or with commercially available software, only short durations are required, and the spike rate model accurately predicted left-sided TLE in both our internal and external cohorts, we believe the spike rate model is clinically useful. The accuracy at predicting right-sided TLE was low in the internal cohort, and so we would not recommend using this model to predict right-sided TLE without further validation in larger external cohorts. To promote reproducibility of our results, we provide a free online calculator to apply our model. Clinicians and researchers can input spike rates from NREM sleep in the left and right temporal lobe, and the calculator returns the spike rate AI, along with the predicted probabilities of left-sided and right-sided SOZs derived from the left- and right-sided models. Our goal is for clinicians to be able to use this model, along with other electroclinical data, to help guide surgical planning.

### Conclusion

The results of this study support the hypothesis that interictal abnormalities lateralize to the side of seizure generators in TLE. Furthermore, a simple model using spike rates can accurately predict left-sided SOZs in unseen patients. Given the limitations of using seizures to lateralize seizure generators, we believe our study provides additional motivation to use spike rates to aid in surgical planning in patients with suspected TLE.

## Data Availability

https://penn-cnt.github.io/epilepsy_lateralization/

## Acknowledgments

None

## References

1. Semah F, Picot MC, Adam C, et al. Is the underlying cause of epilepsy a major prognostic factor for recurrence? Neurology. 1998;51(5):1256–1262.

2. Gazzola DM, Thawani S, Agbe-Davies O, Carlson C. Epilepsy monitoring unit length of stay. Epilepsy Behav. 2016;58:102–105.

3. King-Stephens D, Mirro E, Weber PB, et al. Lateralization of mesial temporal lobe epilepsy with chronic ambulatory electrocorticography. Epilepsia. 2015;56(6):959–967.

4. Struck AF, Cole AJ, Cash SS, Westover MB. The number of seizures needed in the EMU. Epilepsia. 2015;56(11):1753–1759.

5. Wieser HG, Blume WT, Fish D, et al. Proposal for a new classification of outcome with respect to epileptic seizures following epilepsy surgery. Epilepsia. 2001;42:282–286.

6. Engel J. Update on surgical treatment of the epilepsies. Neurology. 1993;43(8):609–621.

7. von Ellenrieder N, Peter-Derex L, Gotman J, Frauscher B. SleepSEEG: Automatic sleep scoring using intracranial EEG recordings only. J Neural Eng. Published online April 19, 2022. doi:10.1088/1741-2552/ac6829

8. Conrad EC, Bernabei JM, Sinha N, et al. Addressing spatial bias in intracranial EEG functional connectivity analyses for epilepsy surgical planning. J Neural Eng. 2022;19(5). doi:10.1088/1741-2552/ac90ed

9. Wang Y, Sinha N, Schroeder GM, Ramaraju S. Interictal intracranial electroencephalography for predicting surgical success: The importance of space and time. Epilepsia. 2020;61(7):1417–1426.

10. Conrad EC, Tomlinson SB, Wong JN, et al. Spatial distribution of interictal spikes fluctuates over time and localizes seizure onset. Brain. 2020;143(2):554–569.

11. Lucas A, Cornblath EJ, Sinha N, et al. Resting state functional connectivity demonstrates increased segregation in bilateral temporal lobe epilepsy. Epilepsia. 2023;64(5):1305–1317.

12. Fan L, Li H, Zhuo J, et al. The Human Brainnetome Atlas: A New Brain Atlas Based on Connectional Architecture. Cereb Cortex. 2016;26(8):3508–3526.

13. Ojemann WKS, Scheid BH, Mouchtaris S, et al. Resting-state background features demonstrate multidien cycles in long-term EEG device recordings. medRxiv. Published online July 7, 2023. doi:10.1101/2023.07.05.23291521

14. Klimes P, Cimbalnik J, Brazdil M, et al. NREM sleep is the state of vigilance that best identifies the epileptogenic zone in the interictal electroencephalogram. Epilepsia. 2019;60(12):2404–2415.

15. Conrad EC, Revell AY, Greenblatt AS, et al. Spike patterns surrounding sleep and seizures localize the seizure-onset zone in focal epilepsy. Epilepsia. 2023;64(3):754–768.

16. Grewal SS, Zimmerman RS, Worrell G, et al. Laser ablation for mesial temporal epilepsy: a multi-site, single institutional series. J Neurosurg. Published online July 1, 2018:1–8.

17. Kang JY, Wu C, Tracy J, et al. Laser interstitial thermal therapy for medically intractable mesial temporal lobe epilepsy. Epilepsia. 2016;57(2):325–334.

18. Powell HWR, Parker GJM, Alexander DC, et al. Abnormalities of language networks in temporal lobe epilepsy. Neuroimage. 2007;36(1):209–221.

19. Mayeux R, Brandt J, Rosen J, Benson DF. Interictal memory and language impairment in temporal lobe epilepsy. Neurology. 1980;30(2):120–125.

20. Lucas A, Mouchtaris S, Cornblath EJ, et al. Subcortical functional connectivity gradients in temporal lobe epilepsy. Neuroimage Clin. 2023;38:103418.

21. Lemkaddem A, Daducci A, Kunz N, et al. Connectivity and tissue microstructural alterations in right and left temporal lobe epilepsy revealed by diffusion spectrum imaging. Neuroimage Clin. 2014;5:349–358.

22. Voets NL, Beckmann CF, Cole DM, Hong S, Bernasconi A, Bernasconi N. Structural substrates for resting network disruption in temporal lobe epilepsy. Brain. 2012;135(Pt 8):2350–2357.

23. Voets NL, Bernhardt BC, Kim H, Yoon U, Bernasconi N. Increased temporolimbic cortical folding complexity in temporal lobe epilepsy. Neurology. 2011;76(2):138–144.

24. Trenerry MR, Jack CR Jr, Ivnik RJ, et al. MRI hippocampal volumes and memory function before and after temporal lobectomy. Neurology. 1993;43(9):1800–1805.

25. Bernabei JM, Sinha N, Arnold TC, et al. Normative intracranial EEG maps epileptogenic tissues in focal epilepsy. Brain. 2022;145(6):1949–1961.

26. Taylor PN, Papasavvas CA, Owen TW, et al. Normative brain mapping of interictal intracranial EEG to localize epileptogenic tissue. Brain. 2022;145(3):939–949.

27. Lagarde S, Bénar CG, Wendling F, Bartolomei F. Interictal Functional Connectivity in Focal Refractory Epilepsies Investigated by Intracranial EEG. Brain Connect. 2022;12(10):850–869.

28. Cimbalnik J, Klimes P, Sladky V, et al. Multi-feature localization of epileptic foci from interictal, intracranial EEG. Clin Neurophysiol. 2019;130(10):1945–1953.

29. Travnicek V, Klimes P, Cimbalnik J, et al. Relative entropy is an easy-to-use invasive electroencephalographic biomarker of the epileptogenic zone. Epilepsia. 2023;64(4):962–972.

30. Vollmar C, Stredl I, Heinig M, Noachtar S, Rémi J. Unilateral temporal interictal epileptiform discharges correctly predict the epileptogenic zone in lesional temporal lobe epilepsy. Epilepsia. 2018;59(8):1577–1582.

31. Lee SK, Kim KK, Hong KS, Kim JY, Chung CK. The lateralizing and surgical prognostic value of a single 2-hour EEG in mesial TLE. Seizure. 2000;9(5):336–339.

32. Adachi N, Alarcon G, Binnie CD, Elwes RD, Polkey CE, Reynolds EH. Predictive value of interictal epileptiform discharges during non-REM sleep on scalp EEG recordings for the lateralization of epileptogenesis. Epilepsia. 1998;39(6):628–632.

33. Schulz R, Lüders HO, Hoppe M, Tuxhorn I, May T, Ebner A. Interictal EEG and ictal scalp EEG propagation are highly predictive of surgical outcome in mesial temporal lobe epilepsy. Epilepsia. 2000;41(5):564–570.

34. Cendes F, Li LM, Watson C, Andermann F, Dubeau F, Arnold DL. Is ictal recording mandatory in temporal lobe epilepsy? Not when the interictal electroencephalogram and hippocampal atrophy coincide. Arch Neurol. 2000;57(4):497–500.

