## Supplementary materials for "Interictal intracranial EEG asymmetry lateralizes temporal lobe epilepsy"

### Supplemental Methods

*Intracranial EEG recording*

Inclusion criteria were 1) post-implantation clinical determination of TLE, and 2) bilateral electrode coverage of the temporal lobe structures in a mesial-to-lateral trajectory, where the lowest-numbered contacts on each electrode are in or proximate to the mesial temporal structures, and the higher-numbered contacts are in the lateral temporal neocortex (Fig. 1A). We sought out patients with this electrode coverage because 1) we hypothesized that the interictal network in the bilateral temporal lobes would be most relevant to lateralizing TLE, 2) this electrode trajectory provides broad coverage of the temporal lobe, and 3) these electrode trajectories have standardized placement at both centers, which limits the confounder of anatomical variability in placement. We also excluded patients without seizures during the course of their intracranial EEG evaluation.

Electrode configurations (Ad Tech Medical Instruments, Racine, WI) were cortical strip and grid arrays (HUP: 2.3 mm diameter with 10 mm inter-contact spacing) and depth electrodes (HUP: 1.1 mm diameter with 5 mm inter-contact spacing; MUSC: 0.86mm diameter with 5 mm inter-contact spacing), including stereo-EEG. Sampling rates varied from 512-2048 Hz. Signals were referenced to an electrode distant from the suspected seizure generators.

*Intracranial EEG pre-processing*

We selected a continuous 12-hour period of EEG data beginning at 8 pm on the first full day of recording, chosen to be a time period before most seizures occurred and before substantial medication wean, but also late enough to avoid implant and operative sedation effects. We downsampled each patient's recording to 256 Hz and applied the SleepSEEG algorithm to each patient's 12-hour recording (blind to epileptiform activity) in order to measure the sleep stage [1]. Briefly, the SleepSEEG algorithm extracts oscillatory and non-oscillatory features for each EEG channel, then performs unsupervised clustering of channels based on feature variability, uses a multiclass tree to classify each cluster of channels, and finally combines the results across channels to determine the sleep stage.

We next divided the 12-hour period of EEG into 72 10-minute segments, and then selected a random one-minute segment from each 10-minute segment. This downsampling was performed to reduce computational time. We re-selected a segment if it overlapped with any seizure times to ensure we studied only interictal data (we did not specifically exclude time periods immediately before or after seizures). We next removed any electrodes that were not targeting the temporal lobes in a mesial-to-lateral trajectory, and any electrode contacts that did not have a contralateral pair (e.g., we included LA1, targeting the left amygdala, only if RA1, targeting the right amygdala, was also present). We performed this spatial downsampling because analyzing symmetric electrode pairs limits the confounder of inter-contact distance affecting functional connectivity measurements [2,3]. We removed excluded contacts from analysis prior to performing referencing in order to avoid contaminating the reference signal with that of other electrode contacts (Fig. 1A).

We identified and removed channels with substantial artifacts using an automated algorithm described previously [4]. We next performed three different methods of referencing: machine reference, common average reference, and bipolar reference (in which the reference for a given contact was the higher numbered contact). We examined all three choices of reference because the choice of reference is known to affect connectivity measurements [5], and it remains unclear which provides the most clinically relevant information. We then applied a notch filter to remove power line noise (4th order bandstop IIR filter with half-power frequencies 59 and 61 Hz) and a 0.5-80 Hz bandpass filter (4th order bandpass IIR filter).

*Calculation of interictal EEG features*

We calculated several interictal EEG features, summarized in Table S1. All features were calculated in one-second non-overlapping time windows, and then averaged across all time windows in each one-minute segment. For features that we specify as being calculated across canonical frequency bands, these bands include delta (0.5-4 Hz), theta (4-8 Hz), alpha (8-12 Hz), beta (12-30 Hz), and gamma (30-80 Hz). Features include:

*Spike rate*: The frequency of interictal spikes on a given channel. This was performed using a previously published and validated automated spike detector [6,7]. Each signal was low-pass filtered (6th order 30 Hz Butterworth filter) and high pass filtered (6th order 7 Hz Butterworth filter). Amplitude peaks were detected in the filtered data. Each peak was subjected to the following criteria: (1) minimum absolute height, (2) minimum height relative to the baseline deviation, (3) minimum and maximum duration between adjacent peaks and valleys. Numerical thresholds for each criterion were chosen from visual review of a subset of spikes on several patients, and kept constant for all patients. The timing of the spike peak was defined to be the timing of the maximum amplitude of the signal. As an additional artifact removal step, any spike that did not have a co-occurring spike on another channel within 100 ms was discarded.

*Spike recruitment latency*: The average temporal latency of spikes on a given channel. *Spike sequences* were defined as multiple spikes occurring across two or more channels with at most 100 ms between each spike peak and the next spike peak. The latency of a channel for a given spike sequence was defined to be the time difference between the spike peak on that channel and the spike peak on the earliest spiking channel for that sequence (e.g., the latency would be 0 for the first spike and higher for subsequent spikes). The spike latency for each channel was averaged across all spike sequences involving that channel for each one minute segment. Channels with lower spike latencies tend to have earlier spikes in spike sequences.

*Line length*: The sum of the absolute difference between the EEG signal at adjacent time points, divided by number of samples. A signal with a higher line length would tend to have higher amplitudes and higher frequency activity.

*Bandpower*: The average power in a specific frequency range, calculated across broadband frequency (0.5-80 Hz) and canonical frequency bands.

*Spectral entropy*: The measure of a signal's spectral power distribution, calculated as the Shannon entropy of the normalized power distribution in the frequency domain. Conceptually, a higher spectral entropy signal is one that contains less information. We calculated this using Matlab's *pentropy*.

*Pearson correlation*: The Pearson correlation coefficient between EEG signals on a given channel pair. We took the square of this value to obtain a positive number so that our asymmetry index values would be bounded between -1 and +1.

*Cross correlation*: The absolute value of the Pearson correlation coefficient between EEG signals on a given channel pair when the signals are shifted relative to each other by a time lag. We allowed a maximum time lag to be 200 ms, and found the maximum absolute cross correlation across the time-shifted signals.

*Coherence*: The frequency-specific correlation between EEG signals on a given channel pair. We calculated this using Matlab's mscohere, with a window length equal to the sampling frequency.

*Phase locking value (PLV)*: The phase synchrony between the EEG signals on a given channel pair. We calculated this by filtering the EEG signal on each channel to a given canonical frequency band, applying the Hilbert transform, and obtaining the phase. The PLV between the EEG signals on a pair of channels is then defined as

$$PLV(x,y) = \left| \frac{\sum \quad e^{(i*phase(x) - phase(y))}}{N} \right|$$

Where x is channel x, y is channel y, N is the number of time samples, and i is the imaginary unit.

*Relative entropy*: The difference between the probability distributions of the EEG signals on a given channel pair. We followed the methods of the papers that originally described this measure [8,9]. Briefly, we calculated this by filtering the EEG signal on each channel to a given canonical frequency band, then taking the histogram of the distribution of amplitudes within that signal (10 bins), and then measuring the Kullback–Leibler (KL) divergence between the histograms for the given channel pair. Because the KL divergence is non-symmetric, we defined the relative entropy between channels x and y to be the maximum of $D_{KL}(x||y)$ and $D_{KL}(y||x)$. The KL divergence was occasionally infinite, in which case we defined the relative entropy to be undefined.

We calculated the mean feature across all one-minute time windows, excluding time periods for which the sum of the absolute value of the machine-referenced broadband bandpower was $<10^{-10}$ $\mu V^2$ (implying disconnected electrodes at that time). We separately examined features in sleep and wake given prior work suggesting that sleep-wake rhythms of seizure-risk differ between left and right TLE [10], and given work suggesting that interictal features in sleep better localize seizure generators than features in wakefulness [6,9].

To standardize features, we converted bivariate features to a single univariate feature for each electrode contact, defined as the mean edge weight between that contact and all contacts on the same electrode. We only included those contacts on the same electrode in the average to avoid introducing the confounder of inter-electrode distance on connectivity measurements (the inter-contact distances within an electrode are fixed, however the inter-electrode distance can vary from patient-to-patient). This resulted in a single value for each feature for each electrode contact.

*Details of machine learning algorithm to predict SOZ laterality*

In order to simplify the machine learning problem and to separately examine the ease of predicting left versus right SOZ laterality, we built two separate classifiers: one predicting left-sided SOZ (as opposed to either right *or* bilateral), and one predicting right-sided SOZ (as opposed to left or bilateral). For each classifier, we first performed internal validation on the HUP dataset using leave-one-patient-out cross validation. We performed imputation of missing AI values by re-defining missing AIs in the training and testing set to be the median across the remaining patients in the training set. Missing AIs occurred when all spikes for a choice of reference were unilateral (which resulted in an undefined AI for spike latency), when there were no spikes in sleep for a choice of reference (this occurred in one patient for the bipolar reference, and no patients for either other reference), or in one patient with only odd-numbered electrode contacts, resulting in no channels in a bipolar montage (this occurred in a case in which the patient had more electrode contacts than available amplifier channels). We performed principal component analysis (PCA) on the training data, retaining enough components to explain 95% of the variance in the features. We standardized the data prior to performing PCA by subtracting each AI by the mean across patients and dividing by the standard deviation. We then performed LASSO logistic regression for feature selection and regularization on the remaining principal components. We used the Matlab default value for the regularization constant $\lambda$ (equal to 1/N, where N is the training sample size). The logistic regression model used the principal components to predict SOZ laterality with each patient as a single sample. We applied a misclassification cost inversely weighted by the number of patients in the misclassified class in the training data. After setting this cost function, we selected the default model threshold of 0.5

as the probability above which to assign test cases to the positive class. On the held-out testing patient, we applied the same PCA transformation and LASSO logistic regression model derived from the training data to calculate the probability of the SOZ being left or right. To estimate feature importance, we multiplied the PCA transformation matrix by the classifier feature weights in order to derive standardized weights for the original set of AI features.

We next developed two simpler single-feature models: The first single-feature model included only mean spike rate AI. This model was developed to test whether spike rate asymmetry alone could successfully lateralize TLE. The second simpler model included only *binarized* mean spike rate AI, in which the input to the model 1 if the spike rate AI was positive and 0 if negative. This binarized spike model was meant to approximate one qualitative clinical approach to using spikes, which is to consider which side has more spikes, rather than the quantitative difference between sides. For both single-feature spike models, we performed our primary analysis using spikes detected in the common average reference, and we performed secondary analyses using spikes detected in the bipolar and machine references in order to test the sensitivity of our results to this choice.

### Supplemental results

*Many interictal EEG features are highly correlated, and the choice of reference affects features*
We examined the correlation between interictal EEG features. We first measured inter-feature correlation on an electrode contact-level. To do this, we converted bivariate features to electrode contact-specific univariate features by taking the average edge weight across all other electrode contacts. (For this analysis, we did not restrict the average to contacts only on the same electrode). This yielded a single measure for each patient, feature, electrode contact, and reference. We then calculated the Pearson correlation across electrode contacts between all features and choices of reference, yielding a $N_{\text{features x references}}$ x $N_{\text{features x references}}$ correlation matrix for each patient, where $N_{\text{features x references}}$ is the number of features times the number of references (30 x 3 = 90). Fig. S1A shows the average inter-feature correlation matrix across patients for a single choice of reference (common average), and in sleep. There were often high correlations between different frequency band measurements of the same feature. There were also often high correlations or anti-correlations between different features, such as between coherence and phase-locking value, and between coherence and relative entropy (negative correlation). Spike rates were moderately positively correlated with relative entropy and bandpower. Fig. S1B shows the mean (standard deviation) inter-reference feature correlation across patients for different features. Only the broadband frequency-measured features were considered for this analysis. Correlations varied across features, and the correlation in features between machine reference and common average reference tended to be higher than that between bipolar reference and either other reference. Several correlations were less than 0.5, suggesting that the choice of

reference strongly influences interictal EEG feature calculations, and may have a stronger influence than the choice of connectivity measurement.

We repeated this inter-feature correlation analysis, this time measuring the correlation between the AI of the different features. In this analysis, given that there was a single AI measurement for each patient(rather than each electrode contact), inter-feature Pearson correlations were calculated across patients, rather than across electrode contacts. Similar trends were observed as in the electrode contact-level analysis. Fig. S1C shows the inter-feature AI correlation, again restricting analysis to common average reference. Again, we observed high inter-feature correlation for several features. Fig. S1D shows the inter-reference AI correlation (there are no error bars because there is only a single correlation value across all patients for this analysis). Again, we observed high variability of inter-reference correlations across features.

*Asymmetries in spike rates and relative entropy are distinct across SOZ lateralities - good outcome analysis*

We again compared interictal EEG feature AIs between patients with left-sided, right-sided, and bilateral SOZs, now restricting unilateral patients to those with one-year Engel 1 outcomes. Of the 30 unilateral patients (20 left and 10 right), 15 (9 left and 6 right) underwent surgery and had an Engel 1 outcome. Again, the top-ranked AI features involved spike rates and relative entropy (Fig. S2A). We next compared the set of features that best distinguished left from bilateral SOZs versus right from bilateral SOZs (Fig. S2B). Again, spike and relative entropy features best distinguished left from bilateral SOZs (although none were significant after correcting for the false discovery rate). The best feature set to distinguish right from bilateral was again more heterogeneous (and no features were significant).

*SOZ laterality prediction - good outcome analysis*

We again tested the ability of interictal features to predict SOZ laterality in unseen patients, now restricting the unilateral patients in the HUP dataset to be those with Engel 1 surgical outcomes. The AUCs of the ROC of the left- and right-sided internal cross-validation models trained on all features were 0.78 and 0.69, respectively (Fig. S3A). A model trained on only spike rates (CAR reference) achieved better AUCs (0.87 and 0.71 for the left and right models, respectively (Fig. S3B)). Finally, a model trained only on binary spike rates indicating whether there were more spikes on the left or the right performed poorly (AUC of 0.70 and 0.48, respectively (Fig. S3C)). Spike rates, spike timing, and bandpower were the most important features for both the left- and the right-sided models (Fig. 3D).

We further probed the accuracy of the spike-rate only model. Confusion matrices for the left- and right-sided models at the optimal operating points are shown in Figs. S3E and S3F. The balanced accuracy was 74.6% for the model predicting left vs. right/bilateral SOZ, and 62.2% for the model predicting right vs. left/bilateral SOZ. Model accuracies rise quickly with duration sampled, achieving an accuracy similar to the full-duration models with 5 minutes of sampling (Fig. S3G). Finally, we tested how the spike-only models performed in the external MUSC dataset. The balanced accuracies were 72.9% and 78.9% for the left-sided and right-sided models, respectively (Fig. 3H and 3I).

These results were overall similar as those when we include all patients regardless of surgical outcome, although higher AUCs were achieved across all models when we restrict analysis of unilateral patients to those with good outcomes.

**Supplemental Tables**

| **Feature** | **Univariate or Bivariate** | **Frequency** | **Conceptual definition** |
|---|---|---|---|
| Line length | Univariate | Broadband | The sum of the absolute difference between the EEG signal at adjacent time points |
| Bandpower | Univariate | Broadband + canonical | The EEG signal power in a given frequency band |
| Spike rate | Univariate | Broadband | The frequency of interictal spikes |
| Spike recruitment latency | Univariate | Broadband | The latency of an interictal spike in a sequence (where the first spike in a sequence has a latency of 0 ms) |
| Spectral entropy | Univariate | Broadband | The level of uncertainty in an EEG spectral power distribution (how similar the signal is to white noise) |
| Pearson correlation | Bivariate | Broadband | The squared linear correlation between the EEG signals on a pair of electrode contacts |
| Cross correlation | Bivariate | Broadband | The maximum absolute value linear correlation between the time-shifted EEG signals on a pair of electrode contacts |
| Coherence | Bivariate | Broadband + canonical | The frequency-specific correlation between the EEG signals on a pair of electrode contacts |
| Phase | Bivariate | Broadband | The phase synchrony between the EEG signals on |

| | | | |
|---|---|---|---|
| locking value (PLV) | | + canonical | a pair of electrode contacts |
| Relative entropy | Bivariate | Broadband + canonical | The difference between the probability distributions of the EEG signals on a pair of electrode contacts |

Table S1: EEG features. All features studied, their classification as univariate (describing the signal on a single electrode contact) or bivariate (describing the relationship between the signals on a pair of electrode contacts), the frequency ranges over which they were calculated, and their conceptual definitions are shown. Each of the 10 features was calculated across the corresponding number of frequencies shown in the table, and across three choices of reference, yielding a total of (6 single frequency features + 4 multi-frequency features x 6 frequency bands) x 3 = 90 features.

| **Brainnetome parcel label** | **Gyrus** | **Anatomical description** |
|---|---|---|
| STG_L(R)_6_4 | Superior temporal gyrus | Caudal Brodmann area 22 |
| STG_L(R)_6_6 | Superior temporal gyrus | Rostral Brodmann area 22 |
| MTG_L(R)_4_4 | Middle temporal gyrus | Anterior superior temporal sulcus |
| PhG_L(R)_6_1 | Parahippocampal gyrus | Rostral perirhinal cortex |
| PhG_L(R)_6_2 | Parahippocampal gyrus | Caudal perirhinal cortex |
| PhG_L(R)_6_3 | Parahippocampal gyrus | Lateral posterior parahippocampal gyrus |
| PhG_L(R)_6_4 | Parahippocampal gyrus | Entorhinal cortex |
| PhG_L(R)_6_5 | Parahippocampal gyrus | Temporal agranular insular cortex |
| PhG_L(R)_6_6 | Parahippocampal gyrus | Medial posterior parahippocampal gyrus |
| Amyg_L(R)_2_2 | Amygdala | Lateral amygdala |
| Hipp_L(R)_2_1 | Hippocampus | Rostral hippocampus |
| Hipp_L(R)_2_2 | Hippocampus | Caudal hippocampus |

Table S2: Brainnetome parcels included in fMRI connectivity analysis [11].

**Supplemental figures and legends**

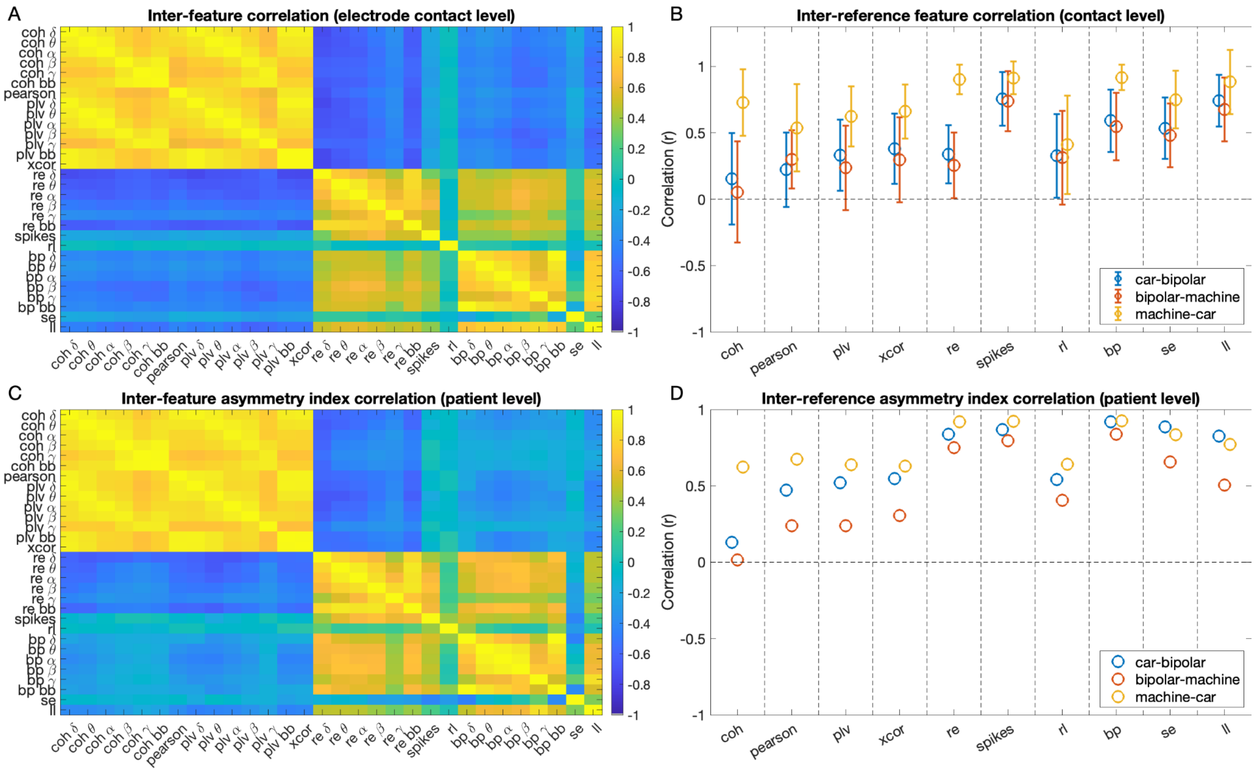


**Figure S1. Inter-feature correlations**. **A:** The correlation between different interictal EEG features across electrodes. All features were calculated using a common average reference and in sleep. Bivariate features were converted to univariate (electrode contact-level) features by taking the average edge weight across all other electrode contact. Several features are highly correlated (bright yellow or dark blue colors indicate high positive and negative correlations, respectively). **B:** The correlation between different reference calculations for EEG features. Each column is a different feature (features are shown for the broadband frequency only). Circles and error bars denote the mean (standard deviation) across patients of the correlation between the features calculated using the denoted pair of references. **C:** The correlation between different interictal feature asymmetry index (AI) values across patients (common average reference). Similar to the result for Fig. S1A, AI values for several features were highly correlated. **D:** The correlation between different reference calculations for AI values. Circles denote the correlated across patients between the AI values calculated using the denoted pair of references. coh = coherence, pearson = pearson correlation, plv = phase-locking value, xcor = cross correlation, re = relative entropy, spikes = spike rates, rl = spike recruitment latency, bp = bandpower, se = spectral entropy, ll = line length. car = common average reference, bipolar = bipolar reference, machine = machine reference. Greek letters denote canonical frequency bands.

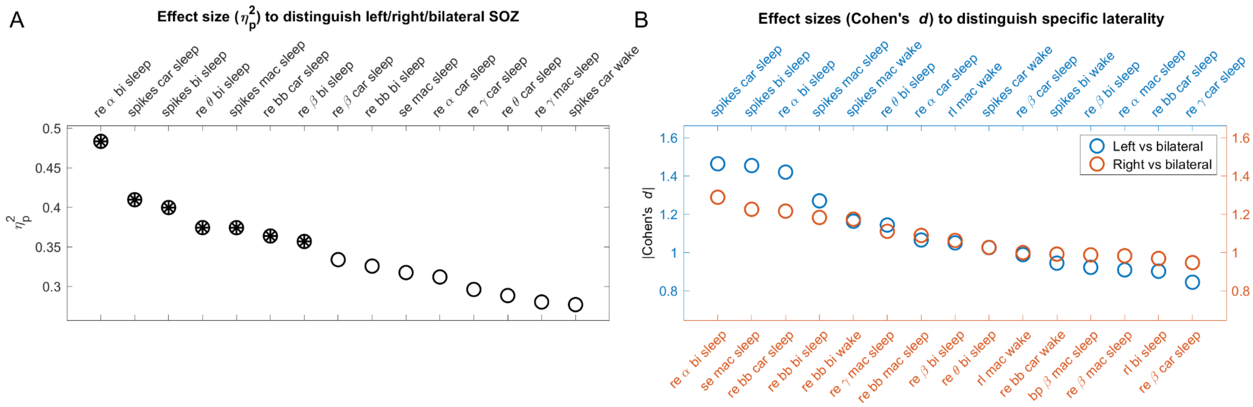


**Figure S2. Comparison of IEEG and fMRI asymmetry index (AI) across seizure onset zone (SOZ) lateralities - good outcome analysis.** Results are from an analysis identical to that for Figure 2A and B, but we removed any unilateral patients who did not undergo surgery or who did not have a one-year Engel outcome of 1.

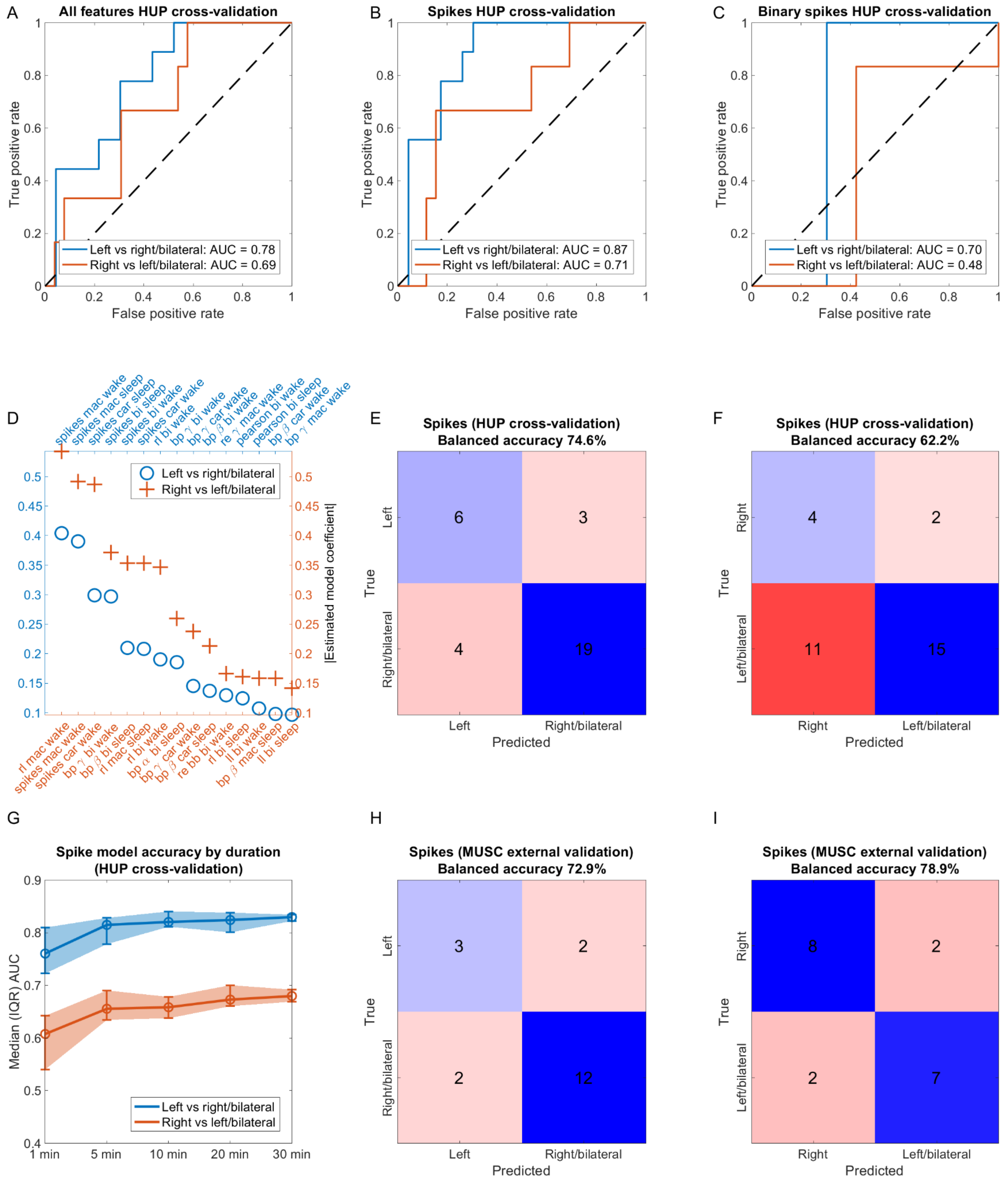


**Figure S3. Classifier to distinguish SOZ lateralities using interictal IEEG asymmetry - good outcome analysis.** Results are from an analysis identical to that for Figure 3, but we restricted the unilateral patients from HUP to be those with one-year Engel surgical outcome of 1.

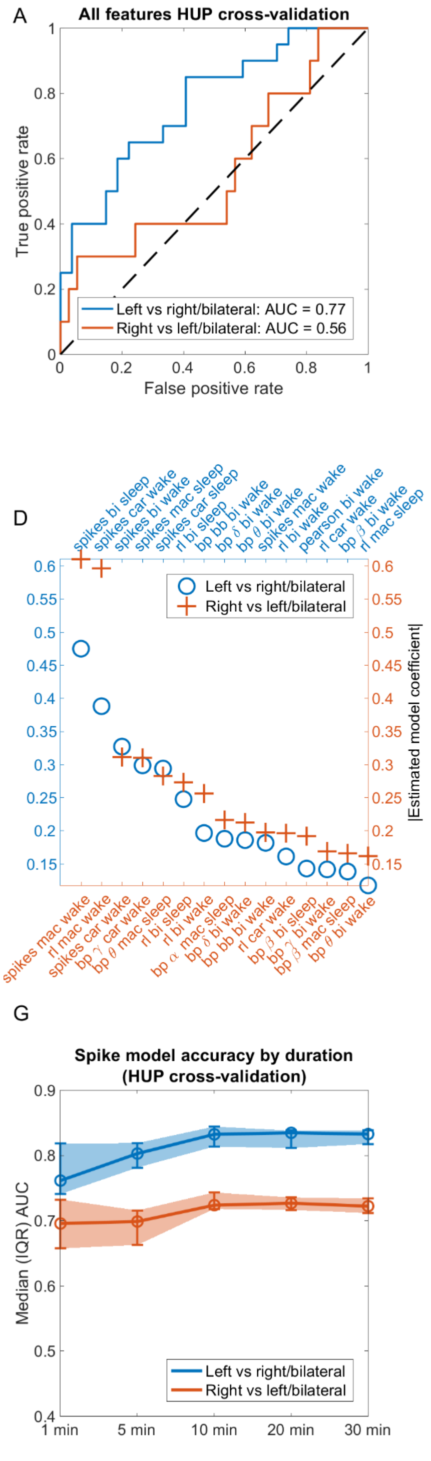


**Figure S4. Classifier to distinguish SOZ lateralities using interictal IEEG asymmetry, where the single feature spike models used spike rates calculated in a bipolar reference.** Results are from an analysis identical to that for Figure 3, but subplots B, C, E, F, G, H, and I (corresponding to single-feature spike models) use spike rates calculated using a bipolar reference.

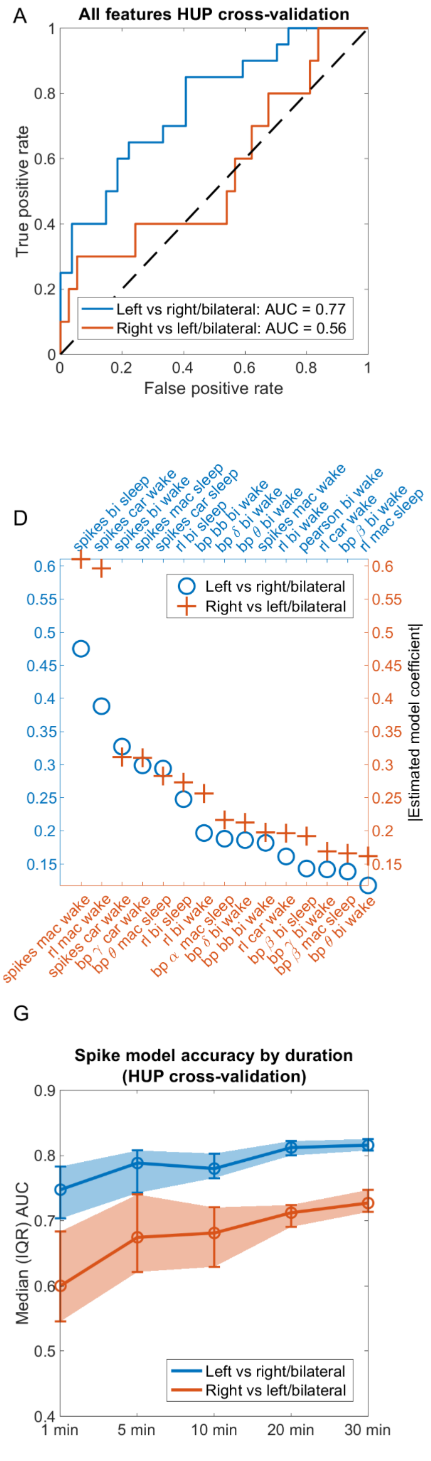

**Figure S5. Classifier to distinguish SOZ lateralities using interictal IEEG asymmetry, where the single feature spike models used spike rates calculated in a machine reference.** Results are from an analysis identical to that for Figure 3, but subplots B, C, E, F, G, H, and I (corresponding to single-feature spike models) use spike rates calculated using a machine reference.

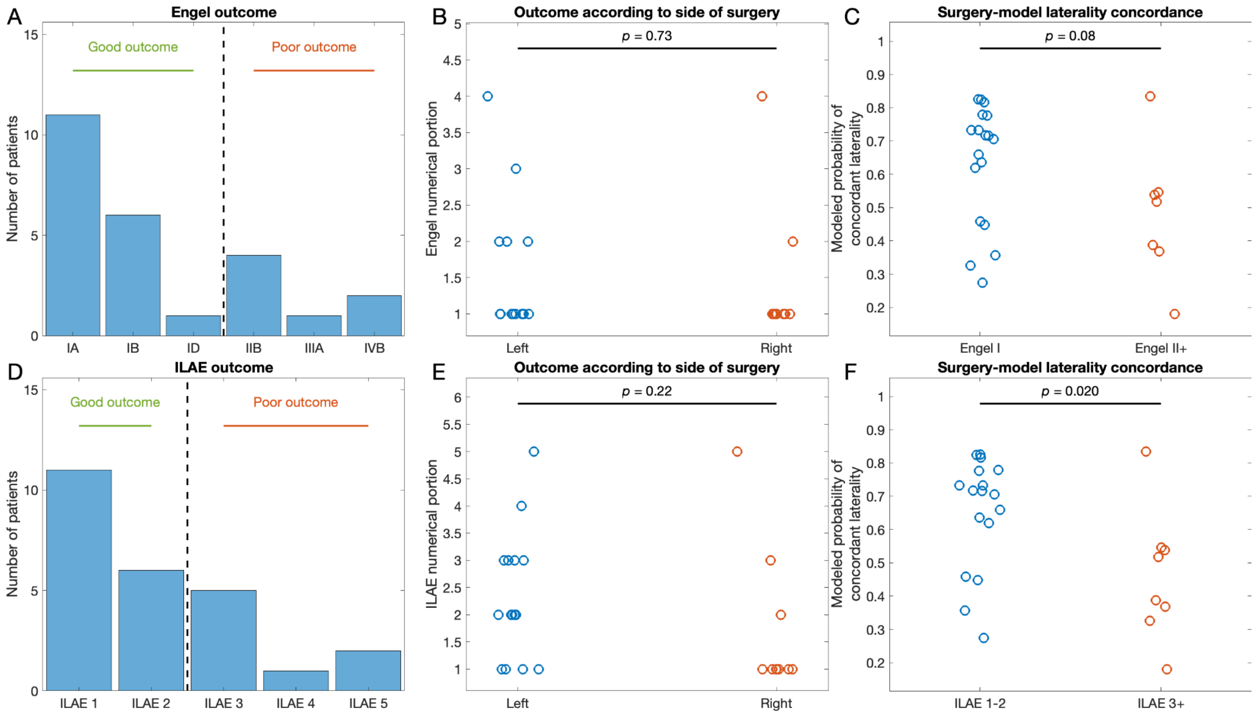


**Figure S6. Surgical outcome prediction using spike rate data calculated with a bipolar reference.** The analysis producing these results is identical to that of the results presented in Figure 4, except that the spike rates used in the model were calculated using a bipolar reference. One patient was excluded from this analysis because they had no spikes in sleep detected in the bipolar referenc.

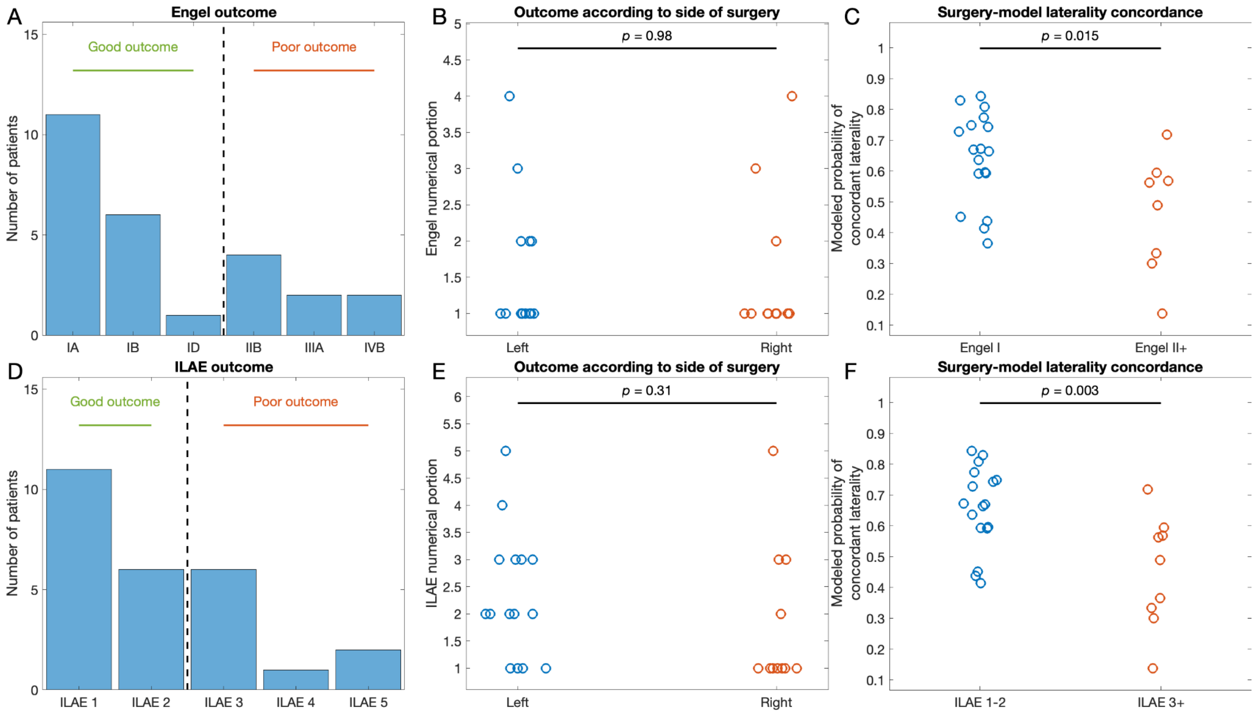

**Figure S7. Surgical outcome prediction using spike rate data calculated with a machine reference.** The analysis producing these results is identical to that of the results presented in Figure 4, except that the spike rates used in the model were calculated using a machine reference.

### References

1. von Ellenrieder N, Peter-Derex L, Gotman J, Frauscher B. SleepSEEG: Automatic sleep scoring using intracranial EEG recordings only. *J Neural Eng*. Published online April 19, 2022. doi:10.1088/1741-2552/ac6829

2. Conrad EC, Bernabei JM, Sinha N, et al. Addressing spatial bias in intracranial EEG functional connectivity analyses for epilepsy surgical planning. *J Neural Eng*. 2022;19(5). doi:10.1088/1741-2552/ac90ed

3. Wang Y, Sinha N, Schroeder GM, Ramaraju S. Interictal intracranial electroencephalography for predicting surgical success: The importance of space and time. *Epilepsia*. 2020;61(7):1417-1426.

4. Conrad EC, Tomlinson SB, Wong JN, et al. Spatial distribution of interictal spikes fluctuates over time and localizes seizure onset. *Brain*. 2020;143(2):554-569.

5. Bastos AM, Schoffelen JM. A Tutorial Review of Functional Connectivity Analysis Methods and Their Interpretational Pitfalls. *Front Syst Neurosci*. 2015;9:175.

6. Conrad EC, Revell AY, Greenblatt AS, et al. Spike patterns surrounding sleep and seizures localize the seizure-onset zone in focal epilepsy. *Epilepsia*. 2023;64(3):754-768.

7. Brown MW 3rd, Porter BE, Dlugos DJ, et al. Comparison of novel computer detectors and human performance for spike detection in intracranial EEG. *Clin Neurophysiol*. 2007;118(8):1744-1752.

8. Travnicek V, Klimes P, Cimbalnik J, et al. Relative entropy is an easy-to-use invasive electroencephalographic biomarker of the epileptogenic zone. *Epilepsia*. 2023;64(4):962-972.

9. Klimes P, Cimbalnik J, Brazdil M, et al. NREM sleep is the state of vigilance that best identifies the epileptogenic zone in the interictal electroencephalogram. *Epilepsia*. 2019;60(12):2404-2415.

10. Latini MF, Oddo S, Anzulovich AC, Kochen S. Daily rhythms in right-sided and left-sided temporal lobe epilepsy. *BMJ Neurology Open*. 2022;4(1):e000264.

11. Fan L, Li H, Zhuo J, et al. The Human Brainnetome Atlas: A New Brain Atlas Based on Connectional Architecture. *Cereb Cortex*. 2016;26(8):3508-3526.
